# Large Language Models for Zero-Shot Procedure Extraction in Orthopedic Surgery: A Comparative Evaluation

**DOI:** 10.1101/2025.08.19.25333995

**Authors:** Ashton Williamson, Nazgol Tavabi, Nishita Kalepalli, Ophelie Lavoie-Gagne, Andre Weiss, Shefali R. Bijwadia, Andrew Sibley, Benjamin Owens, Rafael A. García Andújar, Harsev Singh, Alexandra Santos, Alex Kim, Joseph Murray, Ariana Goli, Leili Sarmadi, Mahad M Hassan, Ata M. Kiapour

## Abstract

**Background:** Operative notes in electronic health records contain critical information for understanding surgical care, yet manual coding is time-consuming, costly, and inconsistent. Large language models (LLMs) promise to transform this process by automatically extracting detailed procedure information — a capability with significant implications for scaling clinical registries and advancing surgical research.

**Methods:** We conducted a large-scale evaluation of state-of-the-art LLMs for zero-shot structured information extraction from orthopedic clinical notes. Fourteen open-source and proprietary models were tested on 800 real operative notes, annotated by both an orthopedic surgeon and an administrator using a curated list of 74 procedure classes. We compared model outputs to human annotations, assessing accuracy and exploring the effects of model scale, reasoning capabilities, and prompt design.

**Results:** Across models, LLMs consistently outperformed administrator-assigned labels, achieving macro-F1 scores above 0.6 and improving over administrative coding by up to 10 points. Larger models and reasoning capabilities further boosted performance, though gains plateaued beyond 30 billion parameters. Performance varied by procedure frequency, revealing clear strengths and persistent challenges for rare or complex cases.

**Conclusion:** Modern LLMs can already outperform routine administrative coding in extracting detailed surgical procedure data, pointing to a future where registry curation could be faster, cheaper, and more consistent. Yet, full alignment with surgical experts remains an open challenge—especially for rare procedures —emphasizing the need for domain adaptation and thoughtful deployment. Our findings illustrate how general-purpose LLMs can advance automated clinical data curation and inform the next generation of surgical informatics.

## Introduction

Clinical registries, also referred to as patient or disease registries, are organized repositories of patient data that are collected through observational means for an express purpose [11]. The construction of clinical registries has emerged as a critical task in healthcare with the potential to enhance clinical research, improve patient quality of care and treatment outcomes, and reduce the cost burden of healthcare [23, 14, 12, 15]. Despite these benefits, the deployment of clinical registries is often hindered by the high costs associated with implementation and maintenance. In a recent survey of organizations operating clinical registries, 64% listed cost as a barrier to sustainability [3] while another survey of high-risk device registries found that only half of respondents received sustainable registry funding [2]. One potential avenue for mitigating the high financial cost of registry construction is the use of secondary data sources, particularly data captured within the electronic health record (EHR) system. While the process of transforming EHR data to fit a registry schema is straightforward for structured data, it is significantly more challenging for unstructured data, like clinical notes. Transforming unstructured clinical notes into variables of interest in the registry is a task that has traditionally necessitated skilled human abstractors, in a process commonly referred to as chart review. Machine-learning-based natural language processing (NLP) algorithms have provided a promising alternative to this paradigm [24, 26, 25, 17], by improving the cost burden, turnaround time, and overall accuracy but are often limited in practice by the need for annotated, in-domain training data.

In this work we investigate the extent to which large language models (LLMs) can assist in the creation and curation of clinical registries by evaluating their ability to extract clinically significant variables from unstructured medical notes in a zero-shot setting. We construct a dataset consisting of operative notes drawn from a high-volume hip-preservation clinic that treats children and young adults, a setting that introduces additional heterogeneity in diagnoses, operative techniques, and documentation styles. We select this domain deliberately because hip surgeries are both common and clinically complex, often involving several different simultaneous procedures. While recent studies have applied large language models to either procedural coding or free-text information extraction, these approaches often fall short for large-scale clinical analysis because raw extracted spans can be inconsistent, ambiguous, or difficult to aggregate. In practice, downstream applications such as registries require consistent, standardized representations of complex procedures. This need is especially acute in domains like orthopedic hip surgery, where common CPT codes may group together distinct operations—such as open femoral head/neck osteochondroplasty, open labral repair, periacetabular osteotomy, and triple innominate osteotomy—under the same code, obscuring important procedural differences [22]. To address this, we framed the task not as open-ended extraction, but as multi-label classification into a carefully defined set of orthopedic procedure classes. This approach bridges the gap between LLMs’ ability to parse rich procedural detail and the practical requirement for structured, analyzable data. Our contributions include the following:

1. We perform the first large-scale evaluation of state-of-the-art large language models (LLMs) for zeroshot multi-label classification of orthopedic operative notes to support clinical registry construction.
2. We develop and utilize an expert-annotated benchmark covering 74 procedure classes across 800 realworld operative notes.
3. We demonstrate that modern LLMs consistently outperform administrator-assigned procedure labels, with the best model improving macro-F1 by 0.10.
4. We analyze the effect of model scaling on performance and efficiency and show diminishing returns beyond 27 billion parameters, while highlighting the benefit of reasoning capabilities (“thinking tokens”) and the impact of different prompting strategies.
5. We leverage our pipeline to construct a comprehensive, 20-year registry of over 80,000 hip procedures across thousands of pediatric patients, which is currently supporting multiple ongoing clinical research efforts.

## Methods

### Dataset

Following IRB approval, we obtained notes from all Boston Children’s Hospital clinics spanning 2000–2023. Using a supervised machine learning pipeline developed in-house [24], we filter out operative notes to include only hip-related operations. This process yielded 30,192 operative notes for 12,203 unique patients across 22,415 encounters. In parallel, with input from a team of orthopedic surgeons, we manually curated a detailed set of common hip procedures with focus on ensuring uniqueness, relevance, and comprehensive coverage, resulting in a final set of 74 distinct procedures (The full list of procedures is provided in Appendix A of Supplementary Materials). Due to the expected long-tail distribution of our labels and the strong effect of sampling on evaluation outcomes [19, 20, 16], we developed a targeted approach (Described in detail in Appendix B of Supplementary Materials) to increase representation of less common classes in the evaluation set. Using this method, we selected 800 operative notes for manual annotation (dataset and sampled-set information, including counts, patient demographics, and note characteristics, are provided in Table 3 of Appendix C in the Supplementary Materials). To assess model alignment with domain experts of varying experience, the sampled notes were annotated once by an orthopedic surgeon and once by an orthopedic administrator with over 10 years of experience, including in conducting chart review. Annotators are each tasked with assigning binary procedure labels to each of the operative notes in the dataset, corresponding to whether the procedure class was conducted as a procedure in the described operation. A visualization of areas of agreement and disagreement across procedure labels between annotators is provided in Figure 4 in Appendix C of Supplementary Materials. All annotations were completed using a custom-built, in-house annotation application developed specifically with this study in mind to facilitate efficient and accurate annotation of operative notes (see Appendix D in Supplementary Materials for details).

### Zero Shot Multilabel Procedure Classification

Given an operative note, the goal is to classify the procedures in the operative note into any of the procedure classes without access to external knowledge sources or reference examples. Models are provided with a description of the task and a list of the procedure classes and prompted to return a JSON object containing a key for each of the procedures and a binary value corresponding to whether a procedure belonging to the class occurred in the operation. Model output is parsed to generate a label vector, where each element corresponds to the presence of a particular procedure from the ordered list of candidate procedures. We experiment with several different open-source LLMs as well as two closed-source, proprietary LLMs (see Table 1). All open-source LLMs are run using the VLLM library on eight NVIDIA H200 GPUs using default sampling parameters suggested by the respective model authors. Proprietary OpenAI models are accessed through a HIPAA-secure institutional API. Models are evaluated against the annotated evaluation set using macro- and micro-averaged Matthew’s Correlation Coefficient, F1, and accuracy. Given the longtail distribution of procedures in our evaluation dataset, we additionally provide an analysis of per-class performance.

**Table 1:** Overview of large language models tested on the hip procedure evaluation dataset. For Mixture of Experts models, we list active parameters, where available. Reasoning Models denote models that support some form of “thinking” tokens to scale test time compute.

| Model | Params | Active Params | Reasoning | Open Weights |
| --- | --- | --- | --- | --- |
| Llama-3.1-8B-Instruct [8] | 8B | - | ✗ | ✓ |
| Llama-3.1-70B-Instruct [8] | 70B | - | ✗ | ✓ |
| Llama-3.1-405b-Instruct [8] | 405B | - | ✗ | ✓ |
| medgemma-4b-it [21] | 4B | - | ✗ | ✓ |
| medgemma-27b-text-it [21] | 27B | - | ✗ | ✓ |
| Qwen3-30B-A3B [29] | 30B | 3B | ✓ | ✓ |
| Qwen3-235B-A22B [29] | 235B | 22B | ✓ | ✓ |
| DeepSeek-R1 [5] | 671B | 37B | ✓ | ✓ |
| DeepSeek-V3-0324 [6] | 671B | 37B | ✗ | ✓ |
| Llama-4-Scout-17B-16E-Instruct [27] | 109B | 17B | ✗ | ✓ |
| Llama-4-Maverick-17B-128E-Instruct [27] | 400B | 17B | ✗ | ✓ |
| gpt-oss-120B [13] | 117B | 5B | ✓ | ✓ |
| gpt-o4-mini-2025-04-16 | - | - | ✓ | ✗ |
| gpt-4o-2024-11-20 | - | - | ✗ | ✗ |

### Model Comparison and Performance Analysis

The proliferation of both open-source and proprietary LLMs and the saturation of many common benchmarks has come to complicate the process of choosing an LLM, particularly in lower-resource settings. To assess the extent to which model architecture and design choices affect performance on our dataset, we evaluate the impact of model size (measured in total model parameters), use of reasoning tokens, and differences in computation time across models. Additionally, given the recent success of reasoning models, a term which we use to refer to models that leverage thinking tokens in their responses [18], we evaluate the extent to which reasoning ability improves performance on our expert-annotated test set. We accomplish this by both measuring the relative performance difference for reasoning models after disabling thinking tokens as well as by comparing performance differences in similarly sized reasoning and non-reasoning models.

### Prompting Strategies

Given the potentially long operative notes and the large number of procedure labels (*>* 70), both the input and expected output for the LLMs can be extensive. Since LLMs have been shown to be susceptible to perturbations in prompt setup [4], we tested different prompting strategies and measured their impact on task performance. The first of these was single-label classification, in which the LLM is provided a full operative note and a single procedure to classify, instead of the full list of procedures. While multilabel classification is a more intuitive approach to the task, it is possible that the complexity introduced by the multilabel prompting setup could adversely affect performance. Providing individual classes, in comparison, may be a more manageable task for an LLM. The second approach was procedure span classification, in which the procedure is extracted from the operative note and provided to the LLM in place of the full text (an example de-identified operative note is provided in Figure 2 of Appendix C in the Supplementary Materials). In effect, this approach both reduces overall input length and reduces the noise introduced by the full body of the operative note, providing a brief clear summary of the operation performed, at the expense of comprehensiveness. Full details of these approaches are provided in the Supplementary Materials Appendices I and J.

## Results

### Dataset

The test set consisted of 800 operative notes, each from a distinct patient and hospital encounter, from 2000 to 2023. Average note length was approximately 1057 tokens (using the GPT-4o tokenizer) with a standard deviation of 913 tokens. In the expert-annotated evaluation set, 90% of the procedure classes (66/74) occur at least once, with an average of 2.64 procedure classes per operative note (*±*2.15) and a maximum of 13 procedure classes assigned to a single note. We calculate Krippendorf’s Alpha for the expert-annotated and administrator-annotated evaluation sets, which we find to be *α* = 0.714.

### Zero Shot Multilabel Procedure Classification

Multilabel classification on the expert annotated set was run across all the models in Table **??**. In order to compare the performance across different models we ranked them in terms of F1-score using a Critical Difference (CD) Diagram [7, 9] on the top 25 most frequent classes, demonstrated in Figure 1. Ranks and significance bars in the CD Diagram are computed using the Friedman test with a post hoc Wilcoxon signed-rank test with Holm’s Correction. Full results of all models across all metrics can be found in Supplementary Materials Appendix F. Based on Figure 1, the best performing model on the expert evaluation set is GPT-o4-mini, achieving an accuracy of 0.986, a macro-averaged F1-score of 0.624, and a macro-averaged Matthews Correlation Coefficient of 0.637. Additionally, GPT-o4-mini outperforms the admin set by 0.09 in micro-averaged F1-score and 0.10 in macro-averaged F1-score. It is also the only LLM that showed a significant difference compared to the administrator annotated labels (*p* = 0.00091), as indicated in the CD diagram in Figure 1. The administrator labels, in contrast, are ranked lower than 12 out of 14 models on the top 25 classes, though they are not significantly different from the other seven worst-performing models.

**Figure 1.**
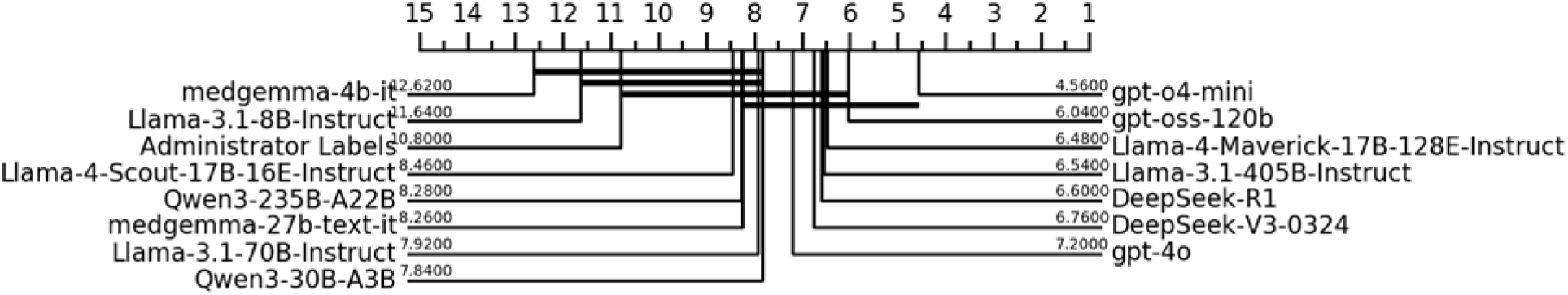
Critical decision diagram ranking evaluated approaches in terms of F1 Score on the top 25 most frequent classes. Horizontal lines denote approaches with no significant difference.

We provide a heatmap of F1-scores across all models and procedure classes in Figure 2. As shown in Figure 2, GPT-o4-mini achieves consistently higher F1-scores across procedure classes. In contrast, the administrator-annotated set shows more variable performance. We additionally compare per-class results in terms of F1-score between the best-performing model on the expert-annotated set and the administrator-annotated set (Table 2 presents the top and bottom 10 procedure classes by frequency, with the full set of results provided in Table 7 in Appendix F of the Supplementary Materials). GPT-o4-mini outperforms the administrator-assigned labels in F1-score for the majority of procedure classes with at least one positive label (43 out of 66). This performance gap is more pronounced among frequently occurring classes —defined as those above the median frequency —where GPT-o4-mini outperforms in 76% of cases, compared to 55% for rarer classes.

**Figure 2.**
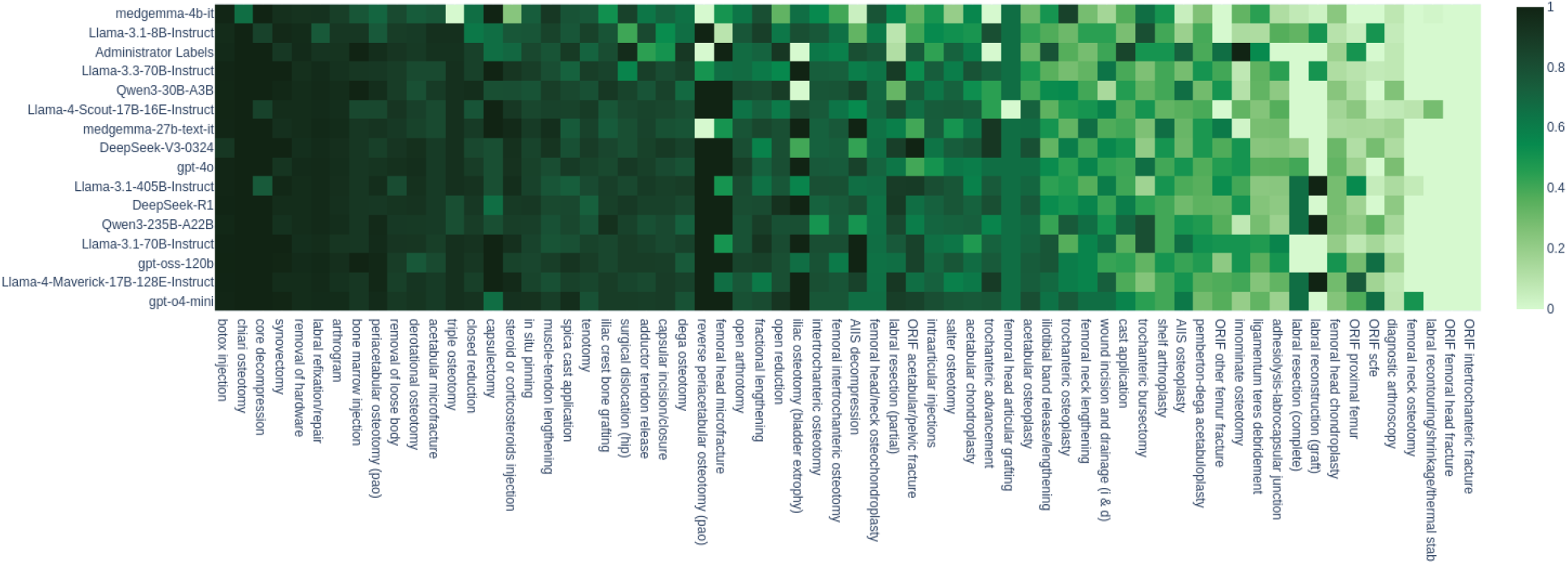
Heatmap of F1-score for each approach evaluated against the expert-annotated set. Procedures are ordered by decreasing mean F1 score.

**Table 2:** Per-class F1-score and Matthew’s Correlation Coefficient for GPT-o4-mini and administrator-assigned labels on the top 10 most common and top 10 least common procedure classes.

|  | Frequency | F1-Score |  | MCC |  |
| --- | --- | --- | --- | --- | --- |
|  |  | GPT-o4-mini | Admin Set | GPT-o4-mini | Admin Set |
| Innominate Osteotomy | 1 | 0.200 | 1.000 | 0.332 | 1.000 |
| ORIF Femoral Head Fracture | 1 | 0.000 | 0.000 | 0.000 | -0.001 |
| ORIF Intertrochanteric Fracture | 1 | 0.000 | 0.000 | -0.001 | 0.000 |
| ORIF SCFE | 1 | 0.667 | 0.000 | 0.707 | -0.002 |
| Labral Reconstruction (graft) | 1 | 0.000 | 0.000 | 0.000 | 0.000 |
| Femoral Neck Osteotomy | 1 | 0.500 | 0.000 | 0.577 | -0.003 |
| Femoral head Microfracture | 1 | 1.000 | 1.000 | 1.000 | 1.000 |
| Shelf Arthroplasty | 1 | 0.400 | 0.500 | 0.499 | 0.577 |
| Reverse PAO | 1 | 1.000 | 0.000 | 1.000 | 0.000 |
| Chiari Osteotomy | 2 | 1.000 | 1.000 | 1.000 | 1.000 |
| Iliac Crest Bone Grafting | 74 | 0.883 | 0.809 | 0.871 | 0.800 |
| Tenotomy | 75 | 0.790 | 0.710 | 0.768 | 0.705 |
| Open Arthrotomy | 75 | 0.793 | 0.677 | 0.778 | 0.669 |
| Arthrogram | 82 | 0.963 | 0.925 | 0.959 | 0.917 |
| Spica Cast Application | 84 | 0.954 | 0.893 | 0.949 | 0.883 |
| Periacetabular Osteotomy | 85 | 0.965 | 0.952 | 0.961 | 0.947 |
| Intraarticular Injections | 86 | 0.805 | 0.435 | 0.784 | 0.435 |
| Removal of Hardware | 128 | 0.959 | 0.965 | 0.952 | 0.959 |
| Botox Injection | 178 | 0.989 | 0.971 | 0.986 | 0.964 |
| Capsular Incision/Closure | 192 | 0.885 | 0.504 | 0.849 | 0.510 |

### Model Comparison and Performance Analysis

As expected, we find an overall positive correlation between model size, measured in total parameters, and micro-averaged F1-score on the expert-annotated evaluation set. We note however, diminishing returns for models larger than 30B parameters (Figure 3). Despite the general trend of higher performance resulting from larger models, we do note some scaling disparity between different families of models. We also observe that models that use thinking tokens perform better than their non-thinking counterparts. In particular, DeepSeek R1 (reasoning) outperforms DeepSeek V3 (non-reasoning) on the expert-annotated set in terms of both micro- and macro-F1 (+0.03 and +0.02, respectively) and GPT-o4-mini outperforms the non-reasoning GPT-4O by 0.02 micro-F1 and 0.04 macro-F1. These improvements, however, fail to account for differences in training setups between the models. For a less biased comparison, we test the Qwen3 reasoning models (Qwen3-235B-A22B and Qwen3-30B-A2B) with and without thinking tokens enabled. We provide the full Reasoning Ablations results in Supplementary Materials Appendix K. This trend holds for the larger of the two models, Qwen3-235B-A22B, which sees a modest decrease in F1 after disabling thinking tokens. In contrast, the smaller model sees negligible performance degradation in terms of micro-F1 (*<* 0.01) and an increase in macro-F1 after disabling thinking tokens. We note, however, that this performance increase comes at a cost of throughput, with reasoning models taking significantly longer to process the full evaluation set, as demonstrated in Figure 3. Furthermore, we observe that models pretrained on biomedical text (MedGemma) outperform identical general-domain models (Gemma), especially smaller ones (see Appendix H), suggesting that augmenting LLM training with in-domain medical text can meaningfully enhance performance on clinical tasks.

**Figure 3.**
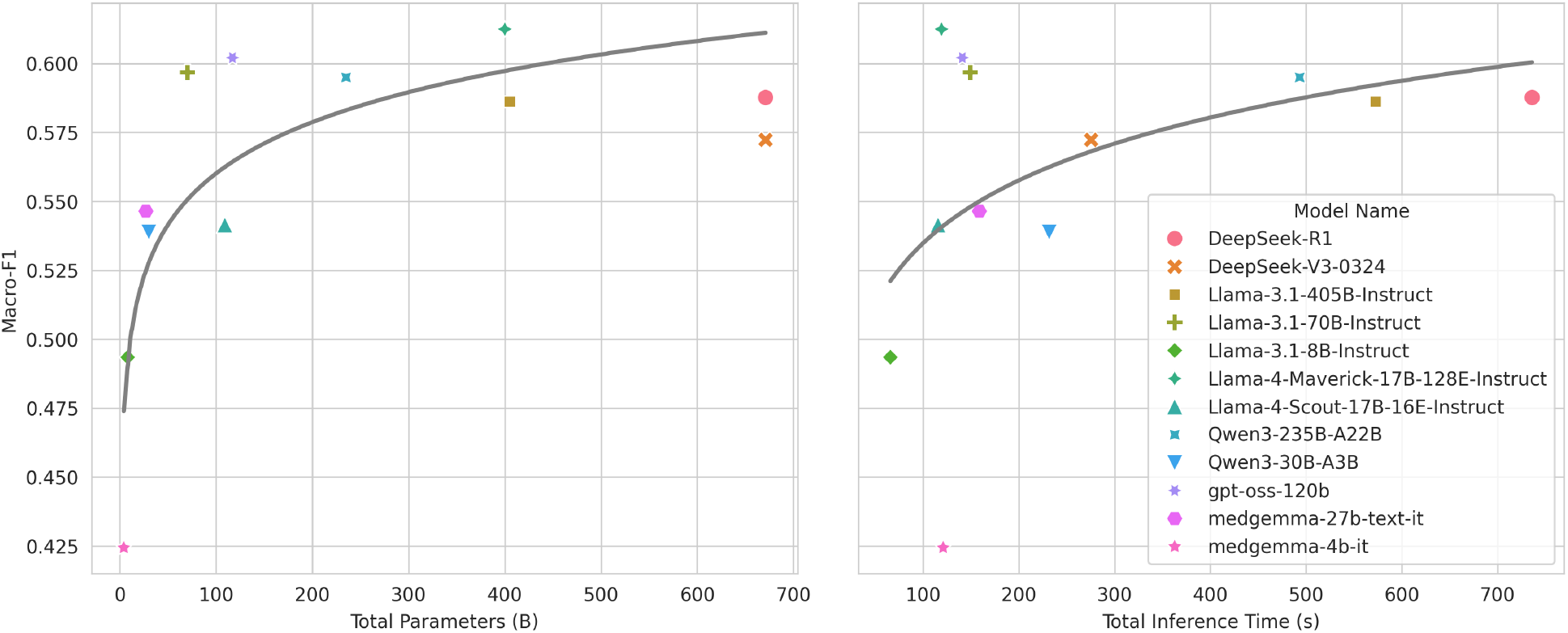
Macro-F1 as a function of total model parameters (in Billions of parameters) and total inference time on the evaluation set (in seconds). Regression lines are created using ordinary least squares in logarithm of x-axis.

### Prompting Strategies

All models evaluated using the single-label classification prompt saw a decrease in micro-averaged F1-score. We also note a general increase in total positive predictions, resulting in a higher micro-averaged recall for all models, though this ultimately outweighed by a comparatively larger decrease in micro-averaged precision. We see similar F1-score performance degradation when classifying extracted procedure sections as opposed to the full note. The exception to this was for the smallest model tested, at 8 billion parameters, which saw a slight increase in F1-score (+3.3%). In contrast to single-label classification, however, all models evaluated on procedure span classification exhibit higher micro-averaged recall but lower micro-averaged precision. The final multi-label classification prompt, its refinement process, and the impact of tailored class instructions on performance are provided in Appendix E.

## Discussion

The extraction of structured data elements from unstructured text within the Electronic Health Record System is a critical task with the potential to enhance the precision of clinical decision-making by enabling large-scale analyses of patient data. While large language models have gained popularity on the task of structured data retrieval in the medical domain [1, 10, 28], the extent to which models can replace expert annotation remains unclear, particularly in specialized sub-domains for which benchmark data is scarce. In this study we investigated the alignment of large language models with human annotators of varying levels of expertise on the task of classifying hip procedures performed in orthopedic hip operations using associated operative notes. Our approach was later used to build a registry of over 20 years of data, capturing 86,555 distinct hip procedures across more than 12,000 patients, with an average age of 14 (See Appendix L in Supplementary Materials for more details). This process completed in under just two hours using GPT-OSS-120B, compared to our human annotators for which the same task took weeks for a dataset of less than 3% of the size.

While GPT-o4-mini demonstrates the best performance on the expert annotated set, the difference be-tween the top-performing models across all procedure classes is not statistically significant and a number of smaller, open-source models perform comparably. Additionally, all approaches demonstrate a high level of per-procedure performance variability, excelling at identifying some procedures while consistently struggling on others. While some of this inter-class performance disparity is model dependent, we find some procedures to be consistently challenging for all LLMs. One example is the set of open reduction and internal fixation (ORIF) procedures, for which the mean macro-F1 across models was less than 0.25. We find that this is most often due to ambiguous references to femoral fracture locations in the notes (which are necessary for classification of 4/7 of these procedures) as well as inconsistent documentation of ORIF procedures by surgeons. Despite the zero-shot nature of the prompting strategy, all models exhibit at least a slight positive correlation between F1-score and procedure frequency in the expert-annotated test set (see Appendix G for a plot of Matthew’s Correlation Coefficient vs Label Frequency). Although we suspect that this discrepancy can be contributed to either shortcomings of the label taxonomy or annotation bias, we do not discard the possibility that the LLMs may have had more exposure to the more common procedures in their training corpus, leading to an increased familiarity with these classes.

While even state-of-the-art models failed to achieve high alignment with expert annotation on the evaluation set, we emphasize the importance of evaluating against annotators with less expertise, given the practical realities of chart review implementation. To this end, we find that state-of-the-art, proprietary LLMs outperform the administrative annotator when evaluating against the expert annotator. Perhaps more surprising, however, is that all but two out of thirteen models outperformed the administrative annotator on the expert set. This includes models with just 27 billion parameters, or approximately 4% of the size of the largest open-source LLM tested, DeepSeek-R1/V3. There are, however, a few key limitations to this study. We first note the relatively small size of the dataset, which limits the generalizability of our results. Additionally, both our expert-annotated set and our administrator-annotated set are annotated by single annotators as opposed to multiple individuals. We suspect that including multiple annotators at the same skill level in a dual-entry setup would provide a more robust and representative dataset. Finally, we note the inherent limitations of assigning a discrete taxonomy to complex and varied medical procedures. We conjecture that more well-defined rules for procedure assignment and more transparent class definitions would lead to better replicability in the annotation process.

## Data Availability

The clinical data that support this research are not publicly available due to privacy or ethical restrictions. The data can be requested following proper IRB and materials transfer agreements. IRB contact

## Acknowledgments

We acknowledge funding support from NVIDIA Applied Research Accelerator, Oracle Research, Children’s Hospital Orthopaedic Surgery Foundation, and the Oberg Family Endowment.

