## Supplementary Materials for "Large Language Models for Zero-Shot Procedure Extraction in Orthopedic Surgery: A Comparative Evaluation"

**D Annotation Application** **11**

**E Prompt Engineering** **12**

**F Unabridged Evaluation Metrics** **14**

**G Assessing Disagreement** **17**

**H Medical LLMs** **18**

**I Procedure Span Classification** **19**

**J Single-Label Classification** **20**

**K Reasoning Ablations** **21**

**L Registry Creation** **22**

### Appendix A Procedure List

Acetabular articular cartilage fixation
Acetabular articular cartilage grafting
Acetabular chondroplasty
Acetabular microfracture
Acetabular osteoplasty
Adductor tendon release
Adhesiolysis-head neck junction
Adhesiolysis-labrocapsular junction
Anterior inferior iliac spine (AIIS) decompression
Anterior inferior iliac spine (AIIS) osteoplasty
Arthrogram
Bone marrow injection
Botox injection
Capsular incision/closure
Capsulectomy
Cast application
Chiari osteotomy
Closed reduction
Core decompression
Dega osteotomy
Derotational osteotomy
Diagnostic arthroscopy
Dunn/modified dunn
Femoral head articular cartilage fixation
Femoral head articular grafting
Femoral head chondroplasty
Femoral head microfracture
Femoral head/neck osteochondroplasty
Femoral intertrochanteric osteotomy
Femoral neck lengthening
Femoral neck osteotomy
Fractional lengthening
Heterotopic ossification (HO) excision
Iliac crest bone grafting
Iliac osteotomy (bladder extrophy)
Iliotibial band release/lengthening
Innominate osteotomy
In situ pinning
Intertrochanteric osteotomy
Intraarticular injections
Labral reconstruction (graft)
Labral recontouring/shrinkage/thermal stab
Labral refixation/repair
Labral resection (complete)
Labral resection (partial)
Ligamentum teres debridement
Ligamentum teres repair
Muscle-tendon lengthening
Open arthrotomy
Open reduction
Open reduction and internal fixation (ORIF) acetabular/pelvic fracture

Open reduction and internal fixation (ORIF) femoral head fracture
Open reduction and internal fixation (ORIF) in situ pinning
Open reduction and internal fixation (ORIF) intertrochanteric fracture
Open reduction and internal fixation (ORIF) other femur fracture
Open reduction and internal fixation (ORIF) proximal femur
Open reduction and internal fixation (ORIF) scfe
Pemberton-dega acetabuloplasty
Periacetabular osteotomy (PAO)
Removal of hardware
Removal of loose body
Reverse periacetabular osteotomy (PAO)
Salter osteotomy
Shelf arthroplasty
Spica cast application
Steroid or corticosteroids injection
Surgical dislocation (hip)
Synovectomy
Tenotomy
Triple osteotomy
Trochanteric advancement
Trochanteric bursectomy
Trochanteric osteoplasty
Wound incision and drainage (I & D)

Figure 1: Complete list of hip-procedure class names.

### Appendix B Sampling Strategy

Given a large corpus of text documents, a set of classes to annotate, and an annotation budget (defined by a maximum number of samples that can be annotated), a natural question that arises is how to sample documents from the corpus such that all classes are represented in the sampled set. This is especially salient when the annotation budget is small and the number of classes is large and unevenly distributed across samples in the corpus. While this problem is ultimately unsolvable without prior knowledge of the assignment of classes, we suspect that if labels are assigned non-adversarially according to semantic features of the text, certain feature-based sampling strategies may outperform others at achieving this aim. To this end, we evaluate different sampling algorithms on existing multi-label datasets, assessing both their deviation from the underlying distribution of classes and their ability to maximize representation of all classes in the sampled set.

Concretely, we denote our document corpus as $\mathcal{D}$, our classes as $C = (c_1, c_2, c_3, \ldots, c_n)$, and the number of samples allowed by our annotation budget as $k$. In the multilabel case, each document in $\mathcal{D}$ is assigned a set of labels for the classes in $\mathbf{C}$ which can be represented by a binary-valued, multi-hot vector of length $n$ ($n = |C|$) where a value of 1 for the $i^{th}$ entry corresponds to a positive label for $c_i$. Thus the entire set of labels for the corpus, $D$ can be represented as a binary valued matrix $\mathcal{L}_D$ of shape $|D| \times |C|$ where each row corresponds to the labels for a document in $D$ and each column corresponds to a class in $C$. We similarly refer to a subset of samples from the corpus as $S$ and its corresponding label matrix as $\mathcal{L}_S$. We use the term *label distribution* of a given label matrix to refer to the normalized frequency distribution given by the column sum of the matrix divided by the count of total positive values (see Equation 1).

$$LD(\mathcal{L}) = \left( \frac{\sum_{i=1}^{|\mathcal{D}|} \mathcal{L}_{i1}}{\|\mathcal{L}\|_0}, \frac{\sum_{i=1}^{|\mathcal{D}|} \mathcal{L}_{i2}}{\|\mathcal{L}\|_0}, \ldots, \frac{\sum_{i=1}^{|\mathcal{D}|} \mathcal{L}_{in}}{\|\mathcal{L}\|_0} \right) \tag{1}$$

Given the preceding formulation, we would thus like to sample a set of documents from $\mathcal{D}$ such that the label distribution of the sampled set remains within some bounds of the underlying label distribution, while simultaneously maximizing the representation of classes in the sampled set. To evaluate the extent to which the label distribution of the sampled set deviates from the label distribution of the full corpus $\mathcal{D}$, we use Kullback-Leibler Divergence, defined as follows:

$$D_{KL}(P\|Q) = \sum_{x \in X} P(x) \log \frac{P(x)}{Q(x)} \tag{2}$$

We use the term *Coverage* to refer to the ratio of number of classes present with at least one positive label in the sampled set to the total number of classes.

We evaluate three different document embedding algorithms–pre-trained language model (PLM) embeddings [11], doc2vec [4], and TF-IDF–and several different sampling strategies, including the Kennard-Stone algorithm [2] and a ridge-leverage score-based sampling method [8] as well as agglomerative, spectral and k-means clustering. For TF-IDF embeddings, we use PCA to reduce the embedding dimensionality to 100. For clustering-based approaches, we partition the embedded documents into clusters and select an equal number of samples from each cluster to reach a total of $k$ samples. For k-means clustering, we additionally experiment with different intra-cluster sampling methods, including sampling with probability proportional to distance from the centroid as well as probability proportional to proximity to the centroid. We provide results for uniform random sampling as a baseline. For approaches that require random sampling–both baseline and clustering based approaches, which randomly sample within clusters–we take the average metrics after running 10 sampling trials. We test these approaches on several open-source multilabel text classification datasets, prioritizing those with long-tail class distributions, including MIMIC-IV (ICD-9 and ICD-10 labels) [1], Reuters-21578 [5] and Amazon Categories [7] (see Table 1 for more details). For tractability reasons, we limit all datasets to 50,000 maximum samples and for clustering based approaches, we limit the number of clusters to 20.

Results for coverage and KL1 Divergence for all datasets for $k = 2000$ can be found in Tables 2 and 3, respectively. Intra-cluster sampling results can be found in Table 4. We find a relative inverse relationship between coverage and KL divergence accounting for sample size; that is to say, approaches that prioritize

| Dataset | # of Documents | # of Classes | Mean Occurrences / Class |
|---|---|---|---|
| MIMIC-IV (ICD-9) | 50,000 | 6,823 | 85.45(±482.6) |
| MIMIC-IV (ICD-10) | 50,000 | 12,382 | 58.47(±421.7) |
| Reuters-21578-ModApte (Train) | 9,603 | 115 | 83.9(±315.7) |
| AmazonCat (Test) | 50,000 | 9,065 | 28.18(±237.5) |

Table 1: Summary statistics of multilabel datasets used for sampling evaluation. Note that for datasets with more than 50,000 samples, we randomly select a subset of 50,000 samples for tractability reasons.

coverage tend to make concessions in terms of divergence from the corpus label distribution. We note, however, that the Coverage to KL Divergence ratio varies across approaches, with certain approaches performing better overall than others. In particular, we find that RLS tends to perform well for both doc2vec and PLM-based embeddings, generally achieving higher coverage than uniform random sampling with a similar amount of KL divergence from the full dataset label distribution. In constrast, Kennard-Stone sampling results in considerably lower coverage while simultaneously demonstrating significantly higher KL divergence. Additionally, we find that sampling outliers within each cluster for k-means tends to results in higher average coverage across datasets, consistently outperforming the baseline uniform random sampling approach.

| Embedding Method | Sampling Method | MIMIC-IV (ICD-9) | MIMIC-IV (ICD-10) | Reuters-21578 (ModApte) | AmazonCat |
|---|---|---|---|---|---|
| Uniform Random Sampling | | 0.378 | 0.290 | 0.719 | 0.242 |
| TF-IDF | K-Means | **0.382** | **0.294** | **0.739** | **0.244** |
| | Spectral | 0.379 | 0.291 | 0.706 | 0.243 |
| | Agglomerative | 0.379 | 0.291 | 0.714 | **0.244** |
| | Kennard-Stone | 0.296 | 0.217 | 0.609 | 0.132 |
| | RLS | 0.380 | 0.287 | 0.713 | 0.228 |
| doc2vec | K-Means | 0.381 | 0.292 | 0.719 | 0.244 |
| | Spectral | 0.381 | 0.294 | 0.730 | 0.240 |
| | Agglomerative | 0.383 | 0.292 | 0.722 | **0.245** |
| | Kennard-Stone | 0.245 | 0.188 | 0.452 | 0.120 |
| | RLS | **0.392** | **0.298** | **0.783** | 0.244 |
| PLM | K-Means | 0.379 | 0.291 | 0.718 | 0.243 |
| | Spectral | 0.380 | 0.292 | 0.730 | **0.246** |
| | Agglomerative | 0.379 | 0.291 | 0.735 | 0.242 |
| | Kennard-Stone | 0.250 | 0.185 | 0.583 | 0.119 |
| | RLS | **0.382** | **0.294** | **0.739** | 0.233 |

Table 2: Coverage scores for $k = 2000$ on four datasets. Sampling methods achieving the highest coverage for each dataset per embedding method are bolded.

| Embedding Method | Sampling Method | MIMIC-IV (ICD-9) | MIMIC-IV (ICD-10) | Reuters-21578 (ModApte) | AmazonCat |
|---|---|---|---|---|---|
| Uniform Random Sampling | | 0.103 | 0.133 | 0.023 | 0.265 |
| TF-IDF | K-Means | 0.104 | 0.133 | 0.023 | 0.264 |
| | Spectral | 0.104 | **0.132** | 0.024 | **0.262** |
| | Agglomerative | **0.103** | 0.133 | **0.022** | 0.264 |
| | Kennard-Stone | 0.535 | 0.694 | 0.122 | 0.870 |
| | RLS | 0.106 | 0.136 | 0.024 | 0.281 |
| doc2vec | K-Means | 0.103 | 0.132 | **0.022** | 0.267 |
| | Spectral | 0.104 | 0.133 | 0.024 | **0.262** |
| | Agglomerative | 0.105 | 0.132 | 0.023 | **0.262** |
| | Kennard-Stone | 0.715 | 0.776 | 0.325 | 0.788 |
| | RLS | **0.101** | **0.127** | 0.088 | 0.323 |
| PLM | K-Means | **0.104** | **0.132** | 0.023 | 0.265 |
| | Spectral | **0.104** | **0.132** | **0.021** | 0.267 |
| | Agglomerative | **0.104** | **0.132** | **0.021** | **0.264** |
| | Kennard-Stone | 0.432 | 0.536 | 0.131 | 0.977 |
| | RLS | 0.106 | 0.136 | 0.023 | 0.261 |

Table 3: KL divergence scores for $k = 2000$ on four datasets. Sampling methods achieving the lowest KL Divergence for each dataset per embedding method are bolded.

| Embedding Method | Sampling Method | MIMIC-IV (ICD-9) | MIMIC-IV (ICD-10) | Reuters-21578 (ModApte) | AmazonCat | Average |
|---|---|---|---|---|---|---|
| Uniform Random Sampling | | 0.378 | 0.290 | 0.719 | 0.242 | 0.407 |
| TF-IDF | Random | 0.380 | 0.292 | 0.717 | 0.246 | 0.409 |
| | Center | 0.378 | 0.292 | 0.727 | 0.245 | 0.411 |
| | Outlier | 0.383 | 0.293 | 0.742 | 0.243 | 0.415 |
| doc2vec | Random | 0.381 | 0.293 | 0.716 | 0.243 | 0.408 |
| | Center | 0.379 | 0.293 | 0.713 | 0.246 | 0.408 |
| | Outlier | 0.381 | 0.292 | 0.736 | 0.242 | 0.413 |
| PLM | Random | 0.376 | 0.291 | 0.730 | 0.244 | 0.410 |
| | Center | 0.381 | 0.292 | 0.718 | 0.243 | 0.408 |
| | Outlier | 0.380 | 0.294 | 0.723 | 0.242 | 0.410 |

Table 4: Coverage results for the different intra-cluster sampling approaches for K-means clustering for each dataset as well as the average result across all four datasets.

### Appendix C Dataset

#### C.1 Operative Note Structure

We provide an example operative note in Figure 2. Although there is some variation in the structure of the note template across our dataset, most operative notes follow a similar template. This typically includes sections for summary details of the operation, most importantly *Pre-Operative Diagnosis*, *Procedure*, *Indications* and *Description of Procedure*. The *Pre-Operative Diagnosis* section includes a free text description of the diagnosis motivating the operation, which may or may not be the same as the post-operative diagnosis. The *Procedure* section provides a high-level overview of the procedures performed in the course of the operation, though we find these to be incomprehensive and inconsistently documented. The *Indications* section provides a more detailed insight into the motivations for the operation. Finally, and most critically, *Description of Procedure* includes a detailed, free-text description of the full operation.

> DATE OF SURGERY: [**Date**]
> SURGEON: [**Surgeon**]
> ANESTHESIA: General
> ESTIMATED BLOOD LOSS: Minimal
> COMPLICATIONS: None
>
> PREOPERATIVE DIAGNOSIS: Right hip labral tear
>
> POSTOPERATIVE DIAGNOSIS: Same
>
> PROCEDURE: Right hip arthroscopy with debridement of labral tear
>
> INDICATIONS: The patient exhibits right hip pain recalcitrant to nonoperative treatment. Preoperative examination and MRI were consistent with labral tear and was felt to be a suitable candidate for arthroscopic management. The risks, benefits and treatment alternatives were discussed with the patient. Informed consent was obtained.
>
> DESCRIPTION OF PROCEDURE: The patient was brought into the operating room and general anesthesia was induced. Preoperative antibiotics were given. The patient was positioned supine on the fracture table. All pressure points were well-padded. A well-padded peroneal post was used. Both feet were padded and placed in traction clamps. The right hip and leg were prepped and draped in the usual sterile fashion. Under fluoroscopic assistance, after traction was applied, anterolateral and posterolateral portals were established with spinal needles followed by guidewires followed by cannulas. The hip joint was entered without difficulty. The 70-degree arthroscope was used. Fluid pump at 60 torr was used. Dilute epinephrine in the irrigant was used. Excellent visualization was obtained. The articular surface of the femoral head was normal. The articular surface of the acetabulum was normal except for the superolateral weight bearing dome were there was some partial thickness chronic chondral loss for an area of approximately 7x7 mm. The fovea was normal. The ligamentum teres was not attached to the femoral head. It was not visualized. There were no loose bodies. The anterior inferior acetabular labrum was normal. The anterior superior labrum was torn to the posterior superior region. This was debrided with the arthroscopic shavers and baskets. The posterior and posterior inferior labrum were normal. The portals were closed with 4-0 Monocryl in an interrupted buried fashion. Marcaine with Depo-Medrol was injected into the joint. The patient tolerated the procedure without complication and was delivered to the recovery room in stable condition. The attending was present for the entire case.
>
> POSTOPERATIVE PLAN: Observation status. Follow-up in two weeks for a clinical check.

Figure 2: Example operative note from the evaluation set.

#### C.2 Summary Statistics

| | Evaluation Set | Full Set |
|---|---|---|
| Samples | 800 | 30,192 |
| Patients | 760 | 12,203 |
| Encounters | 787 | 22,415 |
| Sex Distribution (% Female/Male) † | 56/44 | 58/42 |
| Mean Age at Operation | 14.3(±8.4) | 14.2(±8.4) |
| Mean Words per Note | 730(±658) | 746(±696) |

Table 5: Summary statistics for the evaluation set contrasted against the full set from which it was sampled. † Measured as the percent of operative notes for female versus male patients, as opposed to a percentage of unique patients in the cohort.

#### C.3 Procedure Co-Occurrence

To better understand the distribution of procedures in our dataset, we construct and plot a co-occurrence graph of labeled procedures in the expert-annotated set (See Figure 3). Nodes in the graph correspond to procedure classes and edges correspond to cases where two procedure classes are assigned to the same operative note. Edge weights (widths in the visualization) correspond to the total number of times that two procedure classes are assigned to the same note. We find that this plot reinforces the heterogeneity of procedure classes in the dataset, demonstrating coverage of both routine sub-procedures frequently occurring as part of larger operations as well as more isolated procedures.

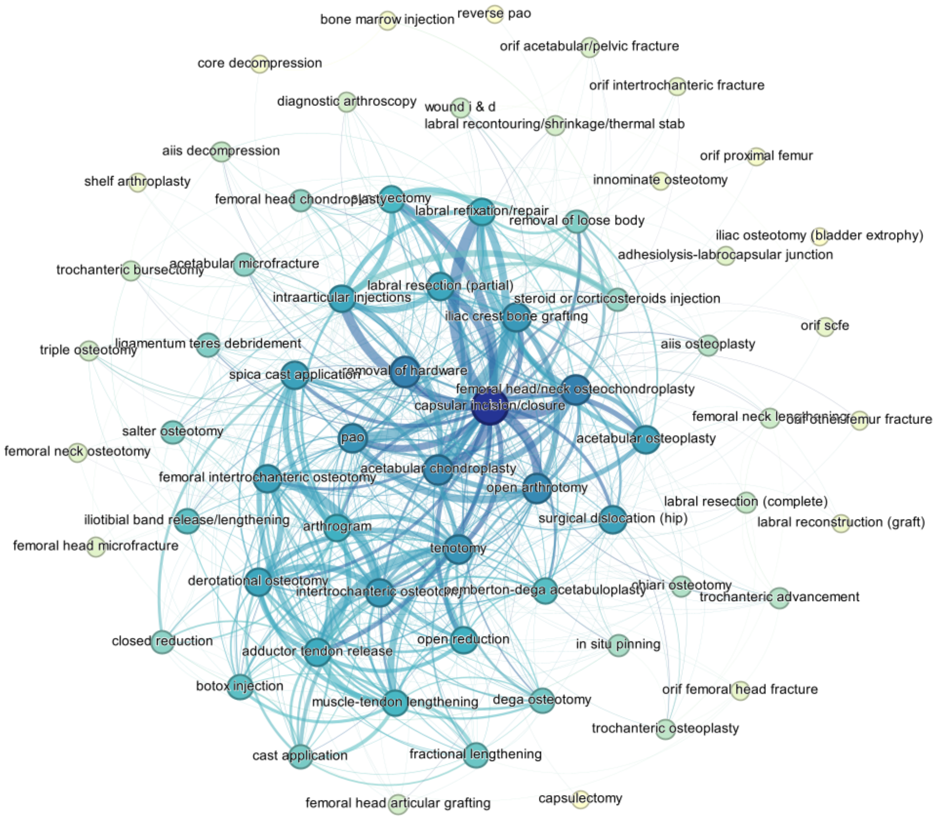

Figure 3: Co-occurrence graph of procedures in our expert-annotated set. Edges correspond to an assignment of the two connected procedures to the same operative note, weighted by frequency of co-assignment.

#### C.4 Annotator Comparison

To better understand cases where annotators disagree, we calculate the Jaccard score per sample for the test dataset. We then generate embeddings for all documents using a small transformer-based language model, which we map to two dimensions using t-SNE [6] and plot, coloring points based on the Jaccard score between the two annotators on the sample, as demonstrated in Figure 4. We identify several distinct clusters and review notes within each of the clusters to evaluate similarities. We find that documents within clusters tend to share a common pre-operative diagnosis–a free text diagnosis description providing by the surgeon (for an example see Figure 2). We label each distinct cluster by these shared pre-operative diagnoses, where possible. The most visually striking cluster is the broader cluster of spasticity cases–spastic diplegia, spastic quadriplegia, and general spasticity–where agreement between annotators is nearly perfect. We attribute this to the fact that these cases also correspond to botox injection treatment, a relatively simple procedure that is both consistently documented and typically performed in isolation, making it easy to classify. In contrast, the closely related labral tear and acetabular impingement clusters show some of the highest rates of disagreement (lowest Jaccard score) across clusters. One explanation for this disagreement is the overall complexity of these operations, which tend to have a large number of sub-procedures despite their short note length. Another possible explanation is the semantic and orthographic similarity of the class names for the procedures involved in these operations, for example the labral tear procedures: labral reconstruction (graft), labral recontouring/shrinkage/thermal stab, labral refixation/repair, labral resection (complete) and labral resection (partial).

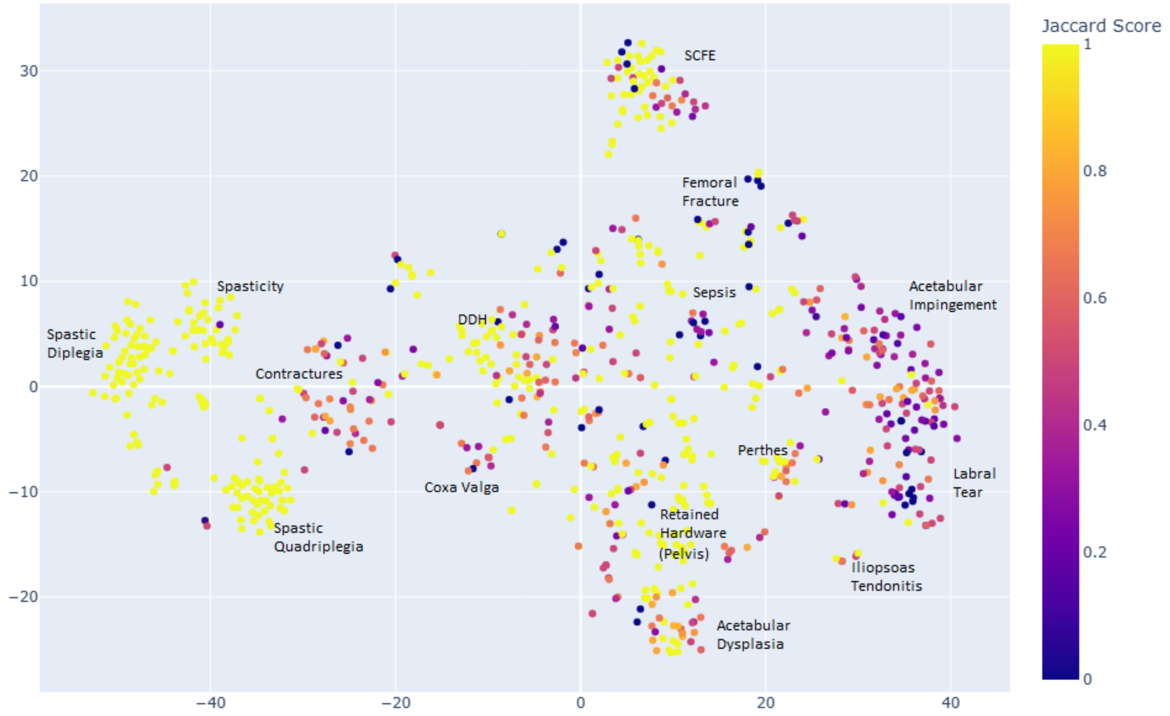

Figure 4: $t$-SNE scatter plot of of operative notes in the evaluation set embedded using a small language model. Points are colored by Jaccard score between the two annotators, with brighter points corresponding to high agreement and darker points corresponding to low agreement. Clusters are labeled qualitatively by common operative diagnosis, as described by the primary surgeon.

### Appendix D Annotation Application

We develop a simple custom web application to assist in the annotation of operative notes. The application is built entirely in python using the Streamlit package and hosted on a secure internal server. The interface provides users with the full text of an operative note and the full set of procedures to annotate 5. Due to the length of operative notes in our set, we choose to display text within a scrolling box so that both note text and procedure classes are visible simultaneously. We provide the full list of procedure selection buttons in the form of a scrollable box, ordered by procedure category, as well as in a searchable dropdown box for easier identification and exploration of the procedure list.

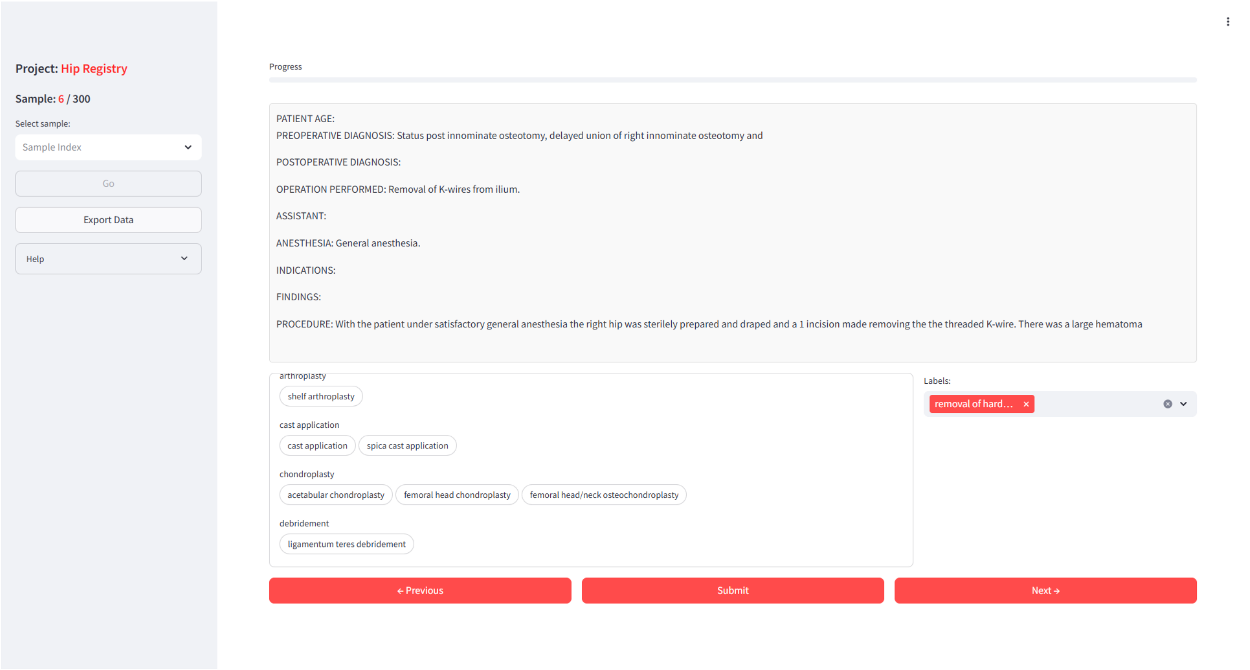

Figure 5: Screenshot of the user interface of the annotation application. A sample (de-identified) operative note is displayed at center, with procedure selection options located directly underneath. The collapsible left pane allows users to check progress and navigate the dataset.

### Appendix E Prompt Engineering

> You are a healthcare professional and you have been given an operative note for a hip-related surgery. Your task is to determine whether any of the following procedures are performed or assigned as a primary procedure:
>
> ⟨List of Procedures⟩
>
> Return your answer as a JSON object with a key for each procedure and a binary value of 0 (procedure absent) or 1 (procedure present). If the procedure is negated in the text (e.g "we chose not to perform...") or the procedure is historical ("patient has recovered from..."), assign it a 0 (not present). Be as precise as possible. All procedure categories should have keys in the final JSON object. Also, note that some procedures may not match any of the provided procedures. Respond only with the JSON object. If you are unsure, simply return an empty JSON object.

Figure 6: Final prompt used for multi-label procedure classification.

The final prompt used for multi-label procedure classification is displayed in Figure 6. The prompt is iteratively refined using a small set of test cases (not included in the evaluation set) to verify both adherence to expected output format and a general understanding of the task definition. To maintain fidelity to a zero-shot approach, we avoid including additional domain knowledge or classification examples. We do, however, perform experiments to evaluate the extent to which more descriptive task instructions and clearer class definitions could improve alignment with our expert annotator. After qualitatively reviewing a subset of LLM mistakes for the most common procedure classes, we select three for which a clear source of confusion could be clearly identified: *diagnostic arthroscopy*, *cast application* and *trochanteric bursectomy*. For the *diagnostic arthroscopy* class, we notice that LLMs seem to fail at distinguishing between *diagnostic* arthroscopy and *surgical* arthroscopies. For the *cast application* class, we find that the low alignment tends to stem from the expert annotator assigning mutually exclusive labels to this class and the *spica cast application* class. LLMs, in comparison, tend to assign both classes when a spica cast application occurs in the operation. Finally, for the *trochanteric bursectomy* class, we notice that the LLMs tend to conflate other bursa procedures (e.g. bursa division/incision) with bursectomies. For each of these cases, we create brief, tailored instructions which we then append to the prompt (see Figure 7).

> **Diagnostic Arthroscopy**: This class is positive only when the arthroscopy is used in a diagnostic capacity. If the arthroscopy is used to perform a procedure, only the performed procedure should be classified as positive (if it belongs to one of the provided classes) and diagnostic arthroscopy should be negative.
>
> **Cast Application**: Apply either this class or spica application but not both. This class should be positive for only non-spica cast applications.
>
> **Trochanteric Bursectomy**: A bursectomy should be referenced as occurring, specifically in the trochanteric region of the femur. Bursa "division" alone does not qualify as a bursectomy.

Figure 7: Additional per-class instructions appended to the prompt for a set of classes with both low alignment with the expert annotator and common, easily-corrected errors. These are not included in the main prompt used to generate classification results (see Figure 6), but are evaluated separately to gauge the ability of bespoke, per-class instructions to reduce common classification errors.

We test both adding one additional class instruction at a time as well as all at once, measuring both the impact on class F-1 Score and micro-averaged F-1 Score, the latter to ensure that error-correction does not lead overall performance degradation. Results are displayed in Table 6. All results are generated using Llama-

4-Maverick-17B-128E-Instruct. We find that adding instructions for Diagnostic Arthroscopy significantly improves performance (0.726 absolute increase in class F-1), while Cast Application instructions lead to only modest performance gains (0.181 absolute increase in class F-1) and Trochanteric Bursectomy instructions lead to no improvement in class F-1, but an overall decrease in micro-averaged F-1 Score. Adding all class instructions simultaneously results in the highest overall micro-averaged F-1 score, but offsets some of the performance gains in the Cast Application cast. In general, results seem to suggest that improved alignment via tailored class instructions is possible to a limited degree, but more review is needed to determine to what extent individual classes might benefit from this approach.

| Prompt | Diagnostic Arthroscopy | Cast Application | Trochanteric Bursectomy | Micro-F1 |
|---|---|---|---|---|
| Baseline | 0.074 | 0.379 | 0.250 | 0.792 |
| Diagnostic Arthroscopy | 0.800 | 0.383 | 0.250 | 0.817 |
| Cast Application | 0.072 | 0.560 | 0.250 | 0.800 |
| Trochanteric Bursectomy | 0.074 | 0.376 | 0.250 | 0.788 |
| All | 0.800 | 0.435 | 0.250 | 0.821 |

Table 6: Classification results (in terms of F-1 Score) for each of the class prompt instructions. *Baseline* corresponds to the standard prompt, without additional class-specific instructions. *All* corresponds to the prompt with all three class-specific instructions appended.

### Appendix F  Unabridged Evaluation Metrics

| | Micro | | | | | |
|---|---|---|---|---|---|---|
| Model Name | Precision | Recall | F-1 | Specificity | MCC | ROC AUC |
| Qwen3-235B-A22B | 0.735 | 0.859 | 0.792 | 0.989 | 0.786 | 0.923 |
| Qwen3-30B-A3B | 0.757 | 0.807 | 0.782 | 0.990 | 0.774 | 0.898 |
| DeepSeek-R1 | 0.768 | 0.865 | 0.813 | 0.990 | 0.808 | 0.927 |
| DeepSeek-V3-0324 | 0.705 | 0.883 | 0.784 | 0.986 | 0.780 | 0.934 |
| medgemma-27b-text-it | 0.669 | 0.863 | 0.754 | 0.984 | 0.750 | 0.923 |
| medgemma-4b-it | 0.409 | 0.816 | 0.545 | 0.956 | 0.558 | 0.885 |
| Llama-3.1-405B-Instruct | 0.677 | 0.923 | 0.781 | 0.984 | 0.782 | 0.953 |
| Llama-3.1-70B-Instruct | 0.669 | 0.901 | 0.768 | 0.983 | 0.767 | 0.941 |
| Llama-3.1-8B-Instruct | 0.476 | 0.884 | 0.619 | 0.964 | 0.632 | 0.922 |
| Llama-4-Maverick-17B-128E-Instruct | 0.692 | 0.925 | 0.792 | 0.985 | 0.792 | 0.954 |
| Llama-4-Scout-17B-16E-Instruct | 0.635 | 0.915 | 0.749 | 0.980 | 0.752 | 0.947 |
| GPT-oss-120b | 0.778 | 0.856 | 0.815 | 0.991 | 0.809 | 0.923 |
| GPT-4o | 0.770 | 0.839 | 0.803 | 0.991 | 0.796 | 0.914 |
| GPT-o4-mini | 0.760 | 0.904 | 0.826 | 0.989 | 0.822 | 0.946 |
| Administrator Labels | 0.839 | 0.657 | 0.737 | 0.995 | 0.734 | 0.826 |

Table 7: Micro-averaged results on the expert-annotated evaluation set.

| | Macro | | | | | |
|---|---|---|---|---|---|---|
| Model Name | Precision | Recall | F-1 | Specificity | MCC | ROC AUC |
| Qwen3-235B-A22B | 0.575 | 0.710 | 0.595 | 0.988 | 0.608 | 0.892 |
| Qwen3-30B-A3B | 0.545 | 0.612 | 0.539 | 0.990 | 0.549 | 0.838 |
| DeepSeek-R1 | 0.579 | 0.672 | 0.588 | 0.990 | 0.598 | 0.871 |
| DeepSeek-V3-0324 | 0.539 | 0.722 | 0.572 | 0.986 | 0.590 | 0.897 |
| medgemma-27b-text-it | 0.530 | 0.679 | 0.546 | 0.984 | 0.562 | 0.872 |
| medgemma-4b-it | 0.409 | 0.617 | 0.424 | 0.956 | 0.443 | 0.822 |
| Llama-3.1-405B-Instruct | 0.546 | 0.769 | 0.586 | 0.983 | 0.608 | 0.922 |
| Llama-3.1-70B-Instruct | 0.568 | 0.720 | 0.597 | 0.983 | 0.609 | 0.894 |
| Llama-3.1-8B-Instruct | 0.451 | 0.724 | 0.493 | 0.962 | 0.517 | 0.885 |
| Llama-4-Maverick-17B-128E-Instruct | 0.580 | 0.747 | 0.612 | 0.984 | 0.627 | 0.910 |
| Llama-4-Scout-17B-16E-Instruct | 0.491 | 0.729 | 0.541 | 0.980 | 0.561 | 0.897 |
| GPT-oss-120b | 0.607 | 0.670 | 0.602 | 0.991 | 0.613 | 0.870 |
| GPT-4o | 0.578 | 0.667 | 0.581 | 0.990 | 0.594 | 0.869 |
| GPT-o4-mini | 0.597 | 0.737 | 0.624 | 0.989 | 0.637 | 0.907 |
| Administrator Labels | 0.609 | 0.510 | 0.525 | 0.995 | 0.533 | 0.783 |

Table 8: Macro-averaged results on the expert-annotated evaluation set.

| Procedure | Label Frequency | GPT-o4-mini | Admin Labels |
|---|---|---|---|
| acetabular articular cartilage fixation | 0 | 0.000 | 0.000 |
| acetabular articular cartilage grafting | 0 | 0.000 | 0.000 |
| acetabular chondroplasty | 67 | 0.754 | 0.612 |
| acetabular microfracture | 8 | 0.889 | 0.933 |
| acetabular osteoplasty | 35 | 0.806 | 0.318 |
| adductor tendon release | 64 | 0.877 | 0.442 |
| adhesiolysis-head neck junction | 0 | 0.000 | 0.000 |
| adhesiolysis-labrocapsular junction | 2 | 0.182 | 0.000 |
| anterior inferior iliac spine (aiis) decompression | 3 | 0.667 | 0.800 |
| anterior inferior iliac spine (aiis) osteoplasty | 5 | 0.625 | 0.714 |
| arthrogram | 82 | 0.963 | 0.925 |
| bone marrow injection | 5 | 0.909 | 1.000 |
| botox injection | 178 | 0.989 | 0.971 |
| capsular incision/closure | 192 | 0.885 | 0.504 |
| capsulectomy | 2 | 0.667 | 0.667 |
| cast application | 32 | 0.571 | 0.847 |
| chiari osteotomy | 2 | 1.000 | 1.000 |
| closed reduction | 20 | 0.930 | 0.865 |
| core decompression | 3 | 1.000 | 1.000 |
| dega osteotomy | 17 | 0.788 | 0.903 |
| derotational osteotomy | 63 | 0.921 | 0.895 |
| diagnostic arthroscopy | 5 | 0.085 | 0.073 |
| dunn/modified dunn | 0 | 0.000 | 0.000 |
| femoral head articular cartilage fixation | 0 | 0.000 | 0.000 |
| femoral head articular grafting | 2 | 0.667 | 0.667 |
| femoral head chondroplasty | 7 | 0.250 | 0.182 |
| femoral head microfracture | 1 | 1.000 | 1.000 |
| femoral head/neck osteochondroplasty | 69 | 0.663 | 0.761 |
| femoral intertrochanteric osteotomy | 50 | 0.769 | 0.590 |
| femoral neck lengthening | 2 | 0.667 | 0.286 |
| femoral neck osteotomy | 1 | 0.500 | 0.000 |
| fractional lengthening | 18 | 0.818 | 0.710 |
| heterotopic ossification (ho) excision | 0 | 0.000 | 0.000 |
| iliac crest bone grafting | 74 | 0.883 | 0.809 |
| iliac osteotomy (bladder extrophy) | 2 | 1.000 | 0.000 |
| iliotibial band release/lengthening | 12 | 0.647 | 0.783 |
| innominate osteotomy | 1 | 0.200 | 1.000 |
| in situ pinning | 38 | 0.932 | 0.831 |
| intertrochanteric osteotomy | 55 | 0.769 | 0.628 |
| intraarticular injections | 86 | 0.805 | 0.435 |
| labral reconstruction (graft) | 1 | 0.000 | 0.000 |
| labral recontouring/shrinkage/thermal stab | 2 | 0.000 | 0.000 |
| labral refixation/repair | 68 | 0.965 | 0.923 |
| labral resection (complete) | 2 | 0.667 | 0.000 |
| labral resection (partial) | 63 | 0.876 | 0.119 |
| ligamentum teres debridement | 6 | 0.333 | 0.545 |
| ligamentum teres repair | 0 | 0.000 | 0.000 |
| muscle-tendon lengthening | 56 | 0.876 | 0.779 |
| open arthrotomy | 75 | 0.793 | 0.677 |
| open reduction | 34 | 0.917 | 0.909 |
| orif acetabular/pelvic fracture | 4 | 0.857 | 0.750 |
| orif femoral head fracture | 1 | 0.000 | 0.000 |

| | | | |
|---|---|---|---|
| orif in situ pinning | 0 | 0.000 | 0.000 |
| orif intertrochanteric fracture | 1 | 0.000 | 0.000 |
| orif other femur fracture | 7 | 0.364 | 0.667 |
| orif proximal femur | 3 | 0.375 | 0.500 |
| orif scfe | 1 | 0.667 | 0.000 |
| pemberton-dega acetabuloplasty | 20 | 0.294 | 0.385 |
| periacetabular osteotomy (pao) | 85 | 0.965 | 0.952 |
| removal of hardware | 128 | 0.959 | 0.965 |
| removal of loose body | 24 | 0.902 | 0.917 |
| reverse periacetabular osteotomy (pao) | 1 | 1.000 | 0.000 |
| salter osteotomy | 8 | 0.769 | 0.714 |
| shelf arthroplasty | 1 | 0.400 | 0.500 |
| spica cast application | 84 | 0.954 | 0.893 |
| steroid or corticosteroids injection | 48 | 0.936 | 0.692 |
| surgical dislocation (hip) | 25 | 0.880 | 0.960 |
| synovectomy | 57 | 0.991 | 0.904 |
| tenotomy | 75 | 0.790 | 0.710 |
| triple osteotomy | 8 | 0.941 | 0.933 |
| trochanteric advancement | 5 | 0.769 | 0.000 |
| trochanteric bursectomy | 2 | 0.444 | 0.500 |
| trochanteric osteoplasty | 4 | 0.750 | 0.364 |
| wound incision and drainage (i & d) | 12 | 0.667 | 0.410 |

Table 9: Per-procedure F-1 score for GPT-o4-mini and administrator-annotated labels, evaluated against the expert annotations. Label frequency represents the number of occurrences of the procedure in the expert-annotated set.

### Appendix G Assessing Disagreement

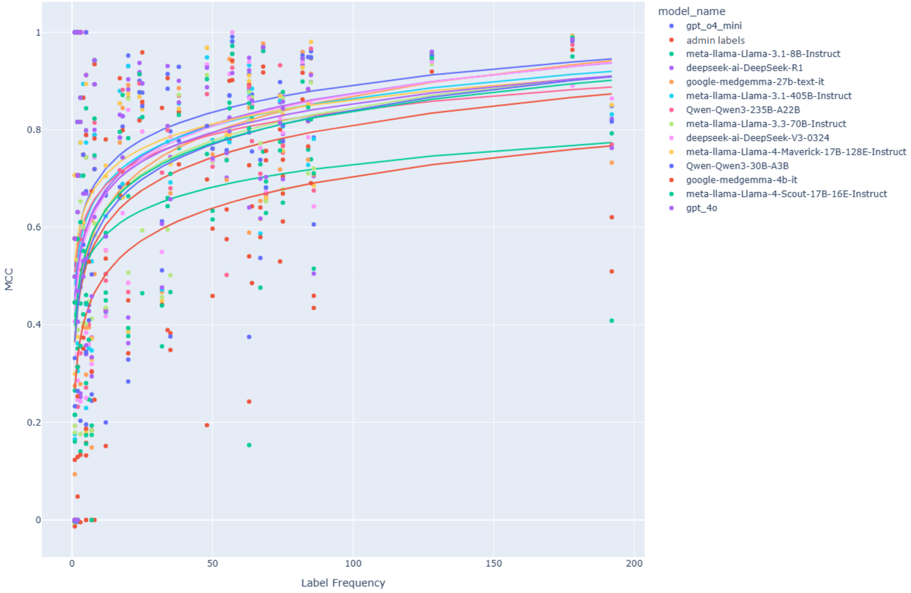

Figure 8: Plot of Matthews Correlation Coefficient vs label frequency in expert-annotated evaluation set.

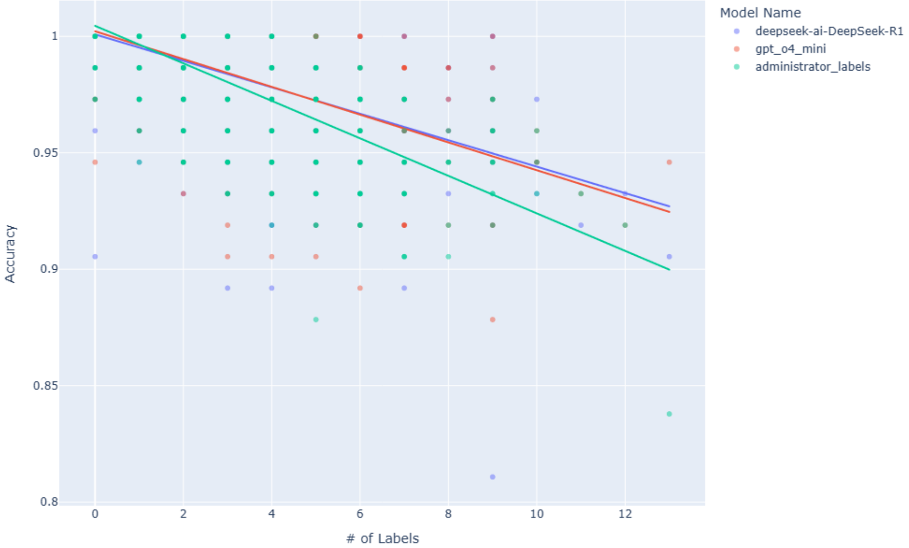

Figure 9: Plot of per-sample accuracy vs number of assigned labels for documents in the expert-annotated evaluation set with regression lines.

| | Pearson | Spearman | Kendall $\tau$ |
|---|---|---|---|
| GPT-o4-mini | 0.36 | 0.13 | 0.07 |
| Administrator Labels | 0.31 | 0.21 | 0.14 |

Table 10: Correlation between per-class F1-score and mean number of co-occurring labels per note for administrator labels and GPT-o4-mini. Mean number of co-occurring labels is calculated by taking the average number of total positive labels assigned across all classes for notes containing a positive label for the target class. High mean co-occurrence suggests that the procedure frequently occurs in tandem with multiple other procedures.

### Appendix H Medical LLMs

Due to the closed-source, proprietary nature of training datasets for many state-of-the-art large language models, it is difficult to determine the amount of medical text to which they have been exposed during training. This ultimately inhibits investigation into the impact of in-domain medical training data on downstream medical language task performance for LLMs. Nonetheless, we seek to assess the extent to which additional exposure to medical text during the training process can improve performance on our evaluation dataset. To this end, we compare two families of models from Google with identical architectures: Gemma [10] and MedGemma [9]. The former, Gemma, is a series of general-purpose, lightweight language models while the latter, MedGemma, is a series of identical models trained predominantly on biomedical data. Results for these models can be found in Table 11. We find a marked improvement in all metrics measured for the MedGemma series, with a much larger relative improvement in the smaller 4 billion parameter model. The largest relative improvement in F1-score for a single procedure class was the for the *removal of loose body* procedure, for which F1 increased by 0.77 for the 4B model and 0.82 for the larger model. These results seem to reinforce the usefulness of augmenting LLM training data with medical text. Although we have no empirical justification for the significant performance increase for the *removal of loose body* class, we suspect that it may have better representation in the training data compared to the other procedures. We leave this avenue of investigation to future work.

| Model | Accuracy | Macro-F1 | Micro-F1 |
|---|---|---|---|
| Gemma-3-4B-it | 0.941 | 0.317 | 0.291 |
| Gemma-3-27B-it | 0.969 | 0.666 | 0.673 |
| MedGemma-4B-it | 0.951 | 0.545 | 0.558 |
| MedGemma-27B-it | 0.980 | 0.754 | 0.750 |

Table 11: Results for both Gemma-3 and MedGemma on the expert-annotated evaluation set.

### Appendix I Procedure Span Classification

Operative notes in our dataset tend to follow pre-defined templates that typically include a section at the top of the note for primary procedures (see the operative note example in Figure 2). In this section, the dictating surgeon includes a free text description of a procedure or set of procedures they identify as central to the operation. While the procedure section may seem like a preferred target for procedure classification as compared to the full text of the operative note, we often find procedure descriptions in this section to be both inconsistently assigned and incomplete. We nonetheless test the extent to which the procedure section can act as a stand-in for the complete operative note by first extracting the text from the procedure section and then running classification on the extracted procedure strings using our standard prompting setup. Results are displayed in Table 12. In terms of micro-averaged F-1, almost all models demonstrate slight performance degradation when classifying extracted procedure headers as opposed to the full note. The exception to this is the smallest model tested–Llama-3.1-8b–with a 3.3 percent relative increase in micro-F1. All models see a decrease in recall score, confirming the expected lack of coverage of the procedure headers. Similarly, all models see a relative increase in precision, since the procedure headers tend to contain short, precise descriptions, whereas the full text of the note introduces more extraneous information, increasing the chance of predicting false positives.

| Model | $\Delta$ Precision (%) | $\Delta$ Recall (%) | $\Delta$ F1 (%) |
|---|---|---|---|
| Qwen3-30B-A3B | 16.3 | −24.0 | −7.4 |
| DeepSeek-R1 | 12.0 | −19.2 | −5.2 |
| medgemma-27b-text-it | 12.3 | −13.9 | −0.9 |
| Llama-3.1-405B-Instruct | 6.8 | −16.2 | −4.3 |
| Llama-3.1-70B-Instruct | 5.1 | −20.2 | −7.4 |
| Llama-3.1-8B-Instruct | 12.5 | −10.3 | 3.3 |
| Llama-4-Maverick-17B-128E-Instruct | 10.5 | −17.4 | −3.5 |

Table 12: Relative percent difference in micro-averaged precision, recall and F1-score on extracted procedure sections instead of the full operative note.

### Appendix J Single-Label Classification

We present the task of hip procedure classification of operative notes as a multi-label classification task, where a classifying agent is required to select a subset of of procedures from a list of candidate procedures. An alternative way of viewing this task is as a binary classification task for each of the 74 procedures. In this setup, the classifying agent is provided with a single procedure class and required to determine its presence in the operation, as described by the operative note (see prompt template in 10). We refer to this setup by the name *single-label*. We evaluate three of the best performing open source large language models (as indicated by multi-label task performance) on the task. Single-label results are displayed in Table 14. We find the single-label approach leads to an increase in recall for all models tested, but a markedly larger decrease precision, resulting in overall lower F1 scores. We suspect that the increase in recall can be attributed to the lack of a reference list of all procedure classes, inhibiting the ability of the LLMs to determine class boundaries. For example, given an operative note that discusses an open reduction and internal fixation procedure for an intertrochanteric fracture and the procedure class *open reduction and internal fixation (ORIF) other femur fracture*, the LLM would be unlikely to abstain from assigning a positive label without knowledge of the existence of the more precise *open reduction and internal fixation (ORIF) intertrochanteric fracture.*

> You are a healthcare professional and you have been given an operative note for a hip-related surgery. Given a category of procedures, your task is to determine whether a procedure belonging to the category occurred as a primary procedure in the operation. Your category is ⟨class_instance⟩.
>
> Your response should be a JSON object with the following schema:
> {
> "⟨class_instance ⟩": 1 if a ⟨class_instance⟩procedure is present in the text else 0
> }
>
> Respond only with the JSON object, no additional text is necessary.

Figure 10: Prompt used for the single-label classification setup.

| Model | Δ Precision (%) | Δ Recall (%) | Δ F1 (%) |
|---|---|---|---|
| DeepSeek-R1 | −21.9 | 10.1 | −9.6 |
| Llama-3.1-405B-Instruct | −10.6 | 1.8 | −5.7 |
| Llama-4-Maverick-17B-128E-Instruct | −22.5 | 0.9 | −14.0 |

Table 13: Relative percent difference in micro-averaged precision, recall and F1-score for single-label classification as opposed to multi-label. Negative values denote a decrease in multi-label performance whereas positive values denote an increase.

Table 14

### Appendix K Reasoning Ablations

| Model | Reasoning | Micro | | | | | |
|---|---|---|---|---|---|---|---|
| | | Precision | Recall | F1 | Specificity | MCC | ROC AUC |
| Qwen3-235B-A22B | On | 0.735 | 0.859 | 0.792 | 0.989 | 0.786 | 0.923 |
| | Off | 0.633 | 0.882 | 0.737 | 0.981 | 0.736 | 0.930 |
| Qwen3-30B-A3B | On | 0.757 | 0.807 | 0.782 | 0.990 | 0.774 | 0.898 |
| | Off | 0.739 | 0.820 | 0.777 | 0.989 | 0.770 | 0.904 |

Table 15: Micro-averaged results for Qwen-3 models with and without reasoning.

| Model | Reasoning | Macro | | | | | |
|---|---|---|---|---|---|---|---|
| | | Precision | Recall | F1 | Specificity | MCC | ROC AUC |
| Qwen3-235B-A22B | On | 0.575 | 0.710 | 0.595 | 0.988 | 0.608 | 0.892 |
| | Off | 0.524 | 0.742 | 0.562 | 0.981 | 0.582 | 0.906 |
| Qwen3-30B-A3B | On | 0.545 | 0.612 | 0.539 | 0.990 | 0.549 | 0.838 |
| | Off | 0.597 | 0.684 | 0.589 | 0.989 | 0.605 | 0.878 |

Table 16: Macro-averaged results for Qwen-3 models with and without reasoning.

### Appendix L Registry Creation

We apply one of our best-performing open-source LLMs, gpt-oss-120b, to the complete set of hip-related operative notes (summary details for which can be found in Table 5). We host the model on-premises utilizing the same compute resources and inference framework that was used for evaluation: 8x H200 GPUs with the VLLM inference engine [3]. Inference on the full 30,192 note set took approximately 2.5 hours total, resulting in 86,555 total procedures. For a comparison of predicted procedure distribution between the full and test sets of hip operative notes, see Figure 11.

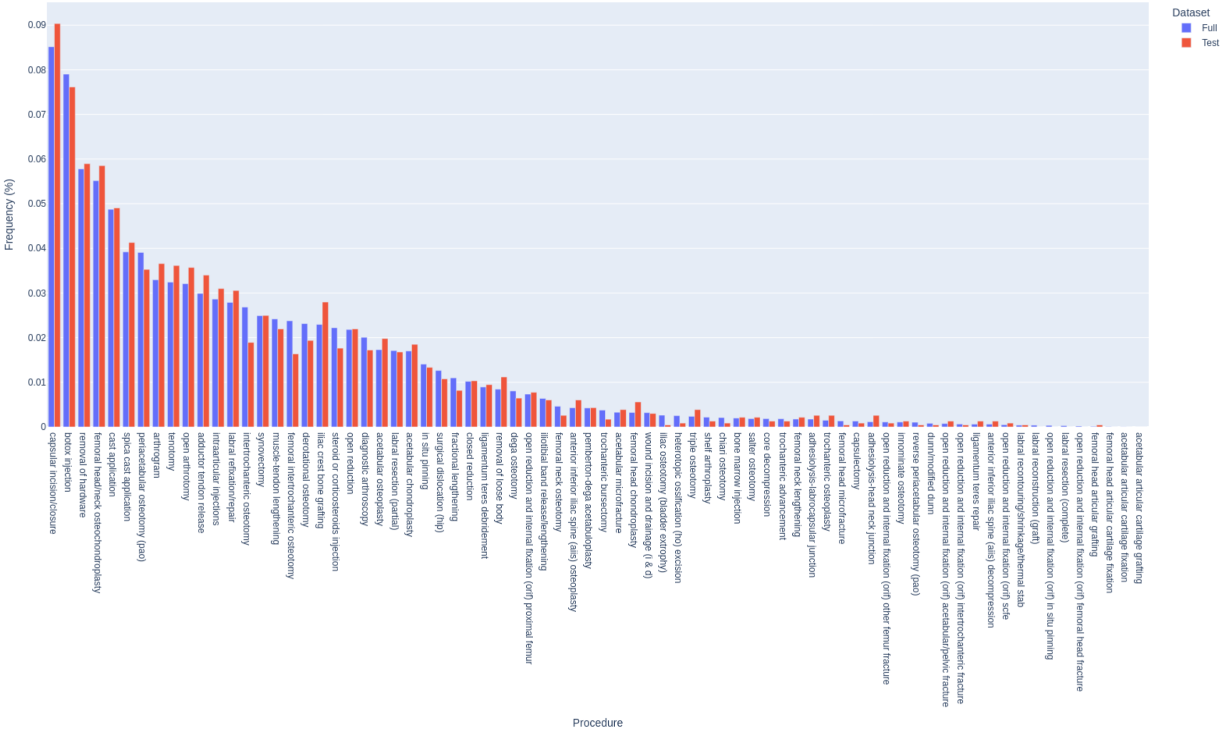

Figure 11: Distribution of predicted procedures in the full dataset vs predicted procedures in the sampled test set, using gpt-oss-120b.
